# Cohort profile: Preliminary experience of 500 COVID-19 postive cases at a South West London District General Hospital

**DOI:** 10.1101/2020.04.28.20075119

**Authors:** R.E Field, I. Afzal, J. Dixon, V.R Patel, P Sarkar, J.E Marsh

## Abstract

This retrospective cohort analysis, reports the demographic data and early outcome of the first 500 patients who were admitted to a District General Hospital in South West London, UK and tested positive to COVID-19. The patients were admitted between 10 January and 10 April 2020; with the first COVID-19 positive diagnosis on 6 March. A surge in admissions started around the 15 March and peaked at the beginning of April.

56.8% of the admissions were male and 43.2% were female. The average age of the 500 admissions was 69.32 years (SD 19.23 years, range 1 week to 99.21 years). By the morning of 14 April 2020, 199 patients had been discharged (Female 89, Male 111), 163 patients had died (female 61, male 102) and 131 remained as in-patients (female 66, male 71).

Fewer than one in twenty deaths occurred in patients below the age of 50 years, in either gender. Mortality rose dramatically, for both genders, after the age of sixty with males being almost twice as vulnerable to dying, as females, during the 7^th^ decade. Males older than their mid-fifties were more likely to die than leave hospital. The same applied to females beyond their mid seventies. We did not see any evidence of a poorer outcome associated with a lower decile for Index of Multiple Deprivation or convincing evidence that any Ethnic minority groups were more likely to die than the White subgroups. When compared to the equivalent medical conditions, normally treated in the early spring, COVID-19 has an increased mortality, adversely affecting more men and an older population.

The mean duration from admission to discharge was 11.29 days (SD 11.50 days). For admission to death, the mean interval was 11.72 days (SD 11.05 days). 62 of the 500 admissions required ventilator support. Of this subgroup, 71% were male and 29% were female. By the morning of the 14 April, no female over the age of 60 had left the intensive care unit alive and no male over the age of 50 had left the intensive care unit alive. At this time-point, 1.2% of the 500 admitted patients had returned alive from the intensive care units, following a period of ventilator support. This figure will rise if prolonged ventilator and renal support proves effective.

While only providing a snapshot of a relatively small number of patients, reviewed over a short time period, from a small geographic area, the data supports the view that the younger members of society are less vulnerable to the adverse sequelae of COVID-19 infection and that any return to normal work and social activities should be considered initially for the individuals who are less than 40-50 years of age. There is an ongoing need for analyses on larger patient cohorts using both demographic and detailed clinical data.

## Manuscript

### Introduction

On 31 December 2019, the Wuhan Municipal Health Commission in Wuhan City, Hubei province, China, reported a cluster of 27 pneumonia cases (including seven severe cases) of unknown aetiology [1]. Ten days later, the China Centre for Disease Control (CDC) reported that a novel coronavirus had been detected as the causative agent for 15 of 59 cases of pneumonia [2]. The following day, the first novel coronavirus genome sequence was made publicly available [3]. The sequence was deposited in the GenBank database (accession number MN908947) and uploaded to the *Global Initiative on Sharing All Influenza Data* (GISAID). A preliminary analysis showed that the novel coronavirus (SARS-CoV-2) clusters with the SARS-related CoV clade and differs from the core genome of known bat coronaviruses.

On 11 February 2020, the World Health Organisation (WHO) Director-General, Dr. Tedros Adhanom Ghebreyesus, announced that the disease caused by this CoV was named COVID-19, coronavirus disease 2019 [4]. In the past twenty years, two previous coronavirus epidemics have occurred, the first started in 2003 and became known as Severe Acute Respiratory Syndrome (SARS)-CoV. SARS-CoV was first recognised in China. The outbreak involved twenty-four countries with over 8,000 cases and 800 deaths. The second was first reported in 2012 and became known as Middle East Respiratory Syndrome (MERS)-CoV. MERS-CoV began in Saudi Arabia with over 2,500 cases and 800 deaths.

On 11 March 2020, the WHO declared COVID-19 as a pandemic and on the evening of 23 March, the UK Prime Minister, Boris Johnson, announced a UK wide lockdown in order to slow the rate of infection of COVID-19 while supporting the stretched NHS. On Thursday 16 April, Foreign Secretary Dominic Raab confirmed that the UK’s lockdown measures would be extended for ‘at least’ another three weeks as the country continued to battle against COVID-19.

On 18 April 2020, in its daily worldwide situation update, the European Centre for Disease Prevention and Control reported 95,247 European deaths related to COVID-19. The five countries reporting most deaths being Italy (22,747), Spain (19,478), France (18,681), United Kingdom (14,576) and Belgium (5,163) [5]. The report recorded that 70,524 cases of COVID-19 had been confirmed in the UK during the preceding two weeks. This equated to a 14-day incidence of 106 cases per 100,000 population. However, the severity of symptoms from COVID-19 infection varies greatly and the number of UK citizens being tested for the virus only exceed 10,000 per day on the 11 April. In consequence, the recorded incidence is likely to be an underestimate.

Within the UK, every NHS hospital has been obliged to halt almost all normal clinical activities including elective operations and elective outpatient appointments. Staff have been re-deployed to support the care of COVID-19 patients. Epsom and St Helier University Hospitals NHS Trust (ESTH) is a large acute trust serving south west London and Surrey. In 2016–17 the Trust provided services to a population of approximately 490,000 people [6]. In this retrospective data analysis, we report on the demographic data and early outcome of the first 500 patients who were admitted to ESTH and tested positive to COVID-19.

### Patients and methods

As part of this retrospective, service evaluation, the first 500 consecutive COVID-19 postive admissions to ESTH were evaluated. All patients included in this service evaluation were swabbed, either as in-patients or when they presented to the emergency department with coronavirus like symptoms. Each swab sample was recorded onto a patient electronic record system with a date and time stamp. If the sample was detected as postive, the COVID-19 postive result was recorded with a date and time stamp.

The data reviewed as part of this service evaluation included: patient demographic data, patient admission data, COVID-19 swab and result data, ventilatory status and ventilation data (if applicable) and outcome data.

The data analysed in this service evaluation was collected from information recorded as part of routine direct clinical care that was undertaken according to agreed treatment plans. Therefore no ethical committee approval was required. No additional contact was made or information was collected from the patient, next of kin, general practitioner and any other health care professional.

The data was extracted from two patient electronic record systems: Clinical Manager (Clinical Manager Version 2.0, vMware Horizon Client. iSOFT) and Ward Watcher software (Critical Care Audit Ltd).

Data was tabulated using Microsoft Excel (Microsoft, Redmond, WA). Data analysis was undertaken on Microsoft Excel (Microsoft, Redmond, WA) and SPSS Version 23 (IBM, SPSS Statistics). Statistical analysis was undertaken using odd ratios, risk ratios and the Fisher’s exact test. The level of significance was set at p= 0.05.

Patient confidentiality and information governance were adhered to throughout this service evaluation and data analysis.

Ethical approval for this service evaluation waived by Epsom & St Helier University Hospitals NHS Trust. All data used in this service evaluation was retrospectively extracted. No patients, Next of Kins or GPs were contacted as part of this service evaluation

Data fields extracted from Clinical Manager:

**Table 1.**

| Field name | Variable type | Comment |
| --- | --- | --- |
| Local patient ID | Continuous |  |
| Gender | Dichotomous | Male or female |
| Date of Birth | Continuous |  |
| Postcode | Text |  |
| Ethnicity | Categorical | Unknown (99), British (A), Irish (B), Any other White background (C), White and Black Caribbean (D), White and Black African (E), White and Asian (F), Any other mixed background (G), Indian (H), Pakistani (J), Bangladeshi (K), Any other Asian background (L), Caribbean (M), African (N), Any other Black background (P), Chinese (R), and Any other ethnic group (S) |
| Admission date | Continuous |  |
| Admission source | Text |  |
| Age at admission | Continuous |  |
| Swab Sample date | Continuous |  |
| Swab Diagnosis date | Continuous |  |
| Discharge date | Continuous |  |
| Discharge location | Text |  |
| Outcome | Categorical | Inpatient, discharged or deceased |
| Date of death | Continuous |  |

Data fields extracted from Ward Watcher:

**Table 2.**

| Field name | Variable type | Comment |
| --- | --- | --- |
| Local patient ID | Continuous |  |
| Date ventilation started | Continuous |  |
| Date ventilation ended | Continuous |  |
| Outcome | Categorical | Still ventilated, have come off the ventilator or deceased |

## Results

The first 500 patients treated at the Epsom & St Helier University Hospitals NHS Trust (ESTH) with a positive diagnosis of COVID-19 infection were admitted to the hospital between the evening of the 10 January 2020 and the early morning of the 10 April 2020. (Figure 1).

**Figure 1.**
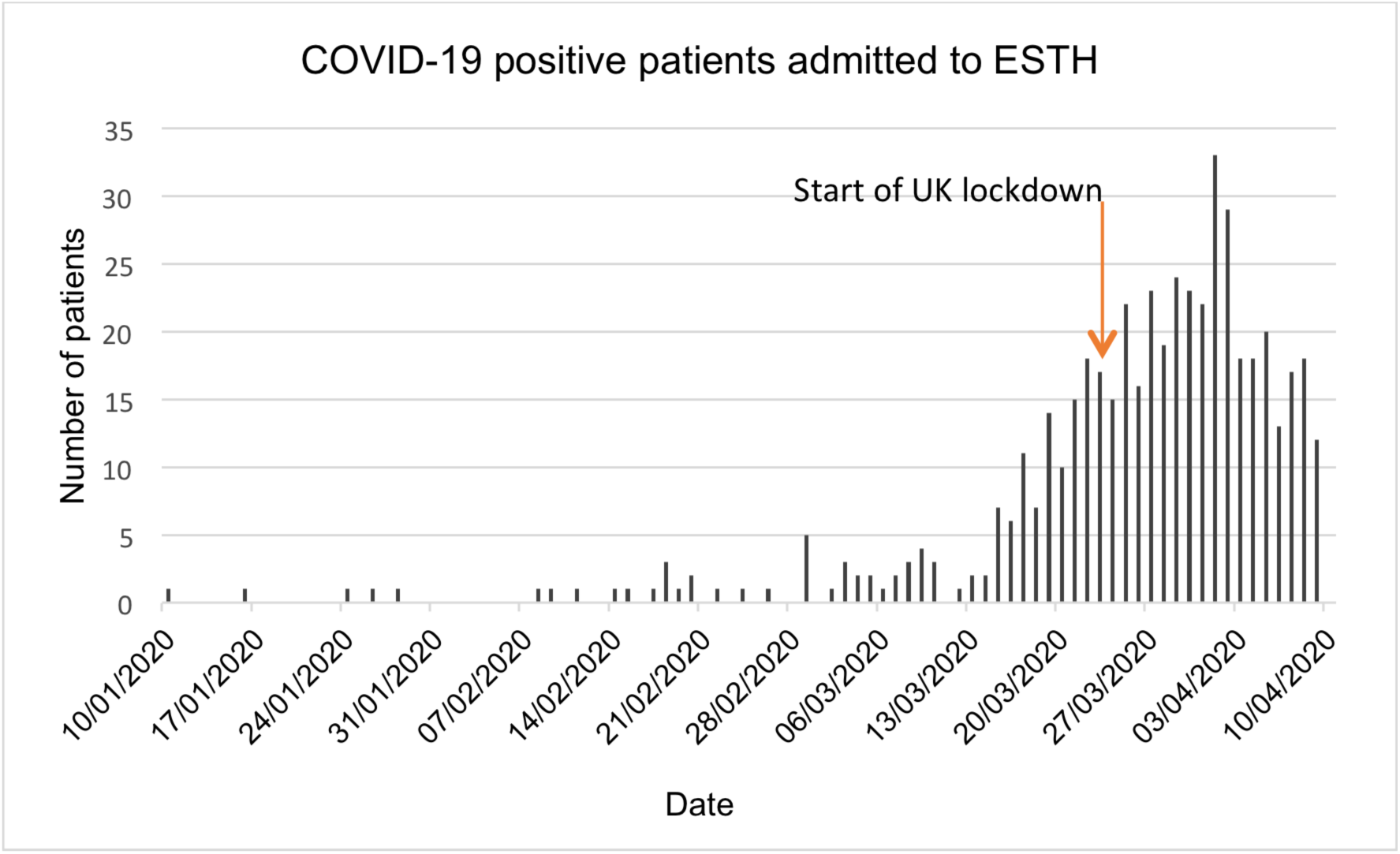

455 of the 500 admissions occurred during the 31-day period from 10 March to 9 April (inc). This equated to an admission rate of 92.86 per 100,000 of the population. A surge in admissions to ESTH started on the 15^th^ of March and peaked just over two weeks later. For the 51 admissions prior to this date, the average interval between admission and testing for COVID-19 was 26.37 days (SD 20.7 days). This interval averaged 1.37 days (SD 3.18 days) for the subsequent 449 admissions.

The average age of the 500 admissions was 69.32 years (SD 19.23 years, range 1 week to 99.21 years). 284 of the admissions were male (56.8%), with an average age of 69.55 years (SD 17.51 years, range 7 weeks to 99.14 years). 216 of the admissions were female (43.2%) with an average age of 68.7 years (SD 21.77 years, range 1 week to 99.21 years).

The profile of the admissions, by age and gender is shown in Figure 2.

**Figure 2.**
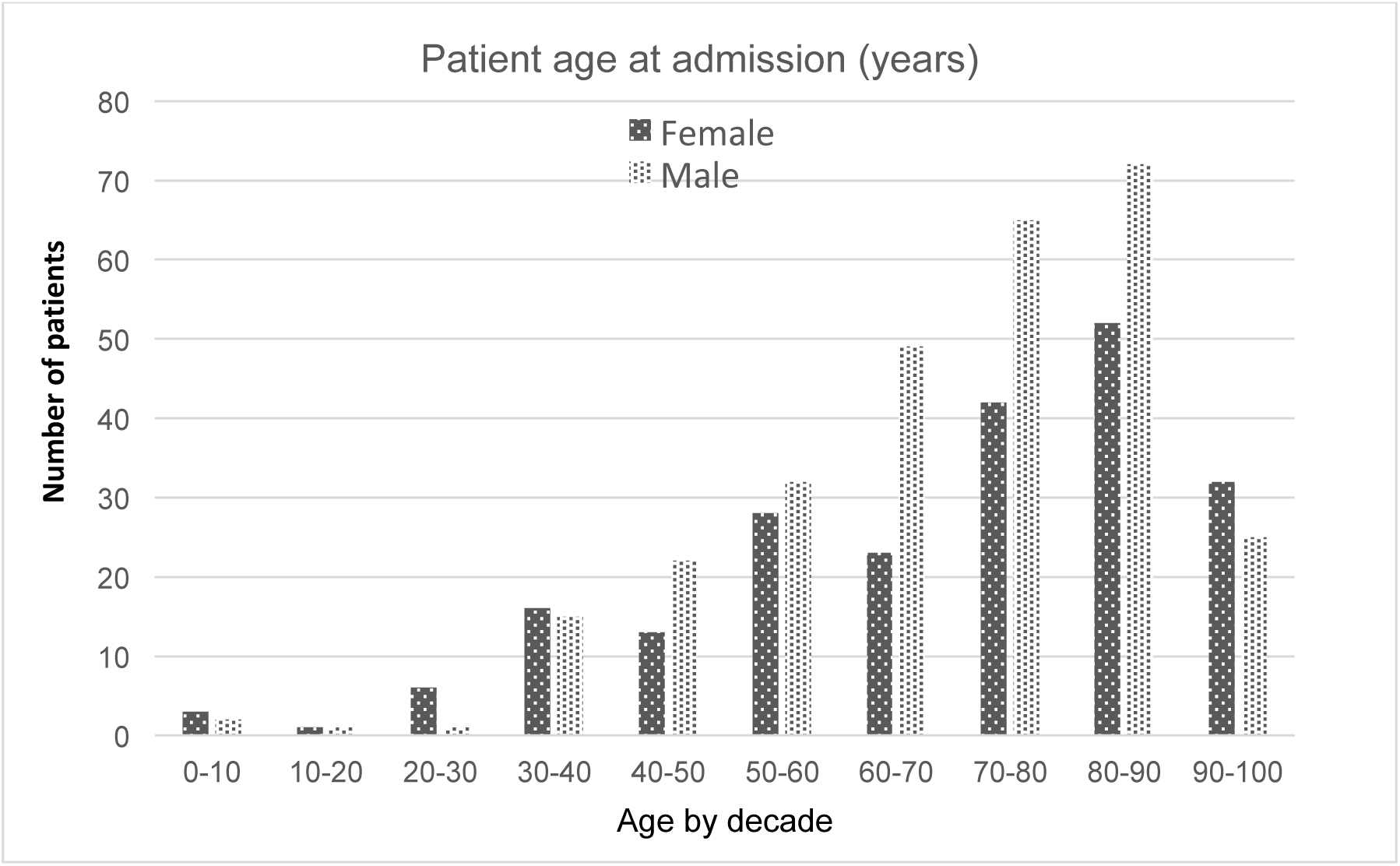

Analysis of the admitted patients, by postcode, for their Index of Multiple Deprivation Decile and the average age of each decile subgroup demonstrated that the patients were predominantly from more affluent homes and that the average age of the patients rose with increasing affluence Figure 3.

**Figure 3.**
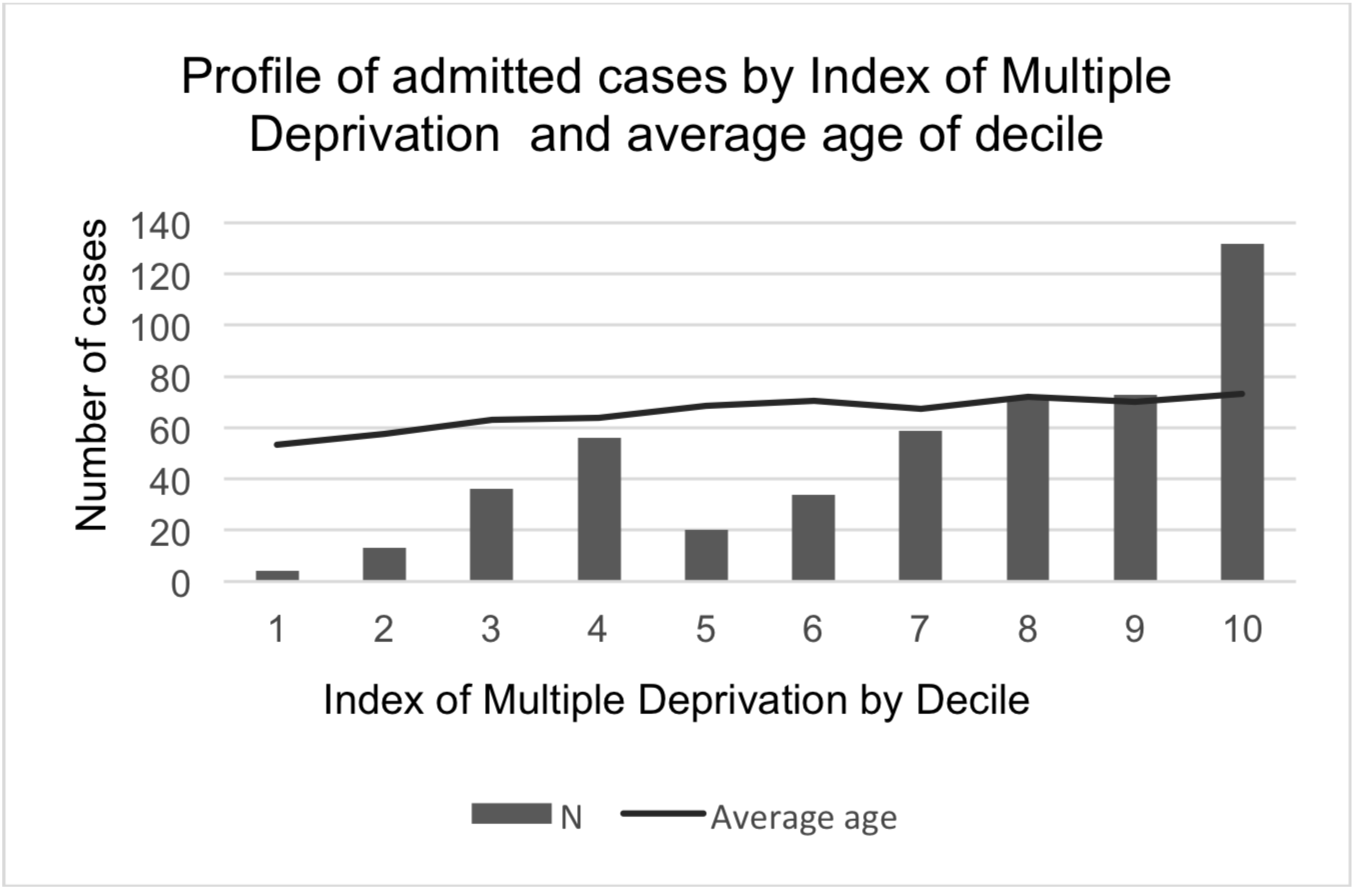

Analysis of the admitted patients was also undertaken by ethnic group. In 34 cases, the ethnic origin was not recorded. 334 cases were recorded as Group A (British), 5 as B (Irish), 26 as C (Any other White background), 4 as F (White and Asian), 3 as G (Any other mixed background), 8 as H (Indian), 8 as J (Pakistani), 2 as K (Bangladeshi), 28 as L (Any other Asian background), 8 as M (Caribbean), 7 as N (African), 11 as P (Any other Black background) and 22 as S (Any other ethnic group). In view of the preponderance of Group A patients, further analysis was undertaken with the patients divided into six groups; Unknown (34), White - A+B+C (365), Mixed race - F+G (7), Asian - H+J+K (46), Black – M+N+P (26) and Other Ethnic – S (22).

By the morning of 14^th^ April 2020, 199 patients had been discharged to home or a care home, one patient had been discharged to a hospice (female 89, male 111). 163 patients had died (female 61, male 102). 131 remained in-patients at ESTH and 6 had been transferred to St George’s or St Thomas’ Hospitals; where they were still in-patients (female 66, male 71).

Further analysis was undertaken to determine whether the outcomes of death or discharge correlated with patient age, gender, deprivation index and ethnicity.

With regard to outcome by age and gender, the number of patients dying or being discharged is shown in Table 3.

**Table 3.**

| **Age range (years)** | **Female died** | **Male died** | **Female discharged** | **Male discharged** |
| --- | --- | --- | --- | --- |
| **0-10** | 0 | 0 | 2 | 2 |
| **10-20** | 0 | 0 | 1 | 1 |
| **20-30** | 1 | 0 | 5 | 1 |
| **30-40** | 1 | 2 | 13 | 9 |
| **40-50** | 0 | 1 | 7 | 19 |
| **50-60** | 3 | 5 | 14 | 15 |
| **60-70** | 8 | 22 | 8 | 14 |
| **70-80** | 10 | 28 | 16 | 18 |
| **80-90** | 19 | 34 | 16 | 21 |
| **90-100** | 19 | 10 | 7 | 11 |
| **Totals** | **61** | **102** | **89** | **111** |

This data is shown by percentage for each subgroup (Figure 4a & b). For both sexes, fewer than one in twenty deaths occurred in patients below the age of 50 years. Mortality rose dramatically for both sexes after the age of sixty with men being almost twice as vulnerable to dying during the 7^th^ decade. The disparity between men and women over the age of 90 is explained by the smaller number of men living to this age.

**Figure 4a.**
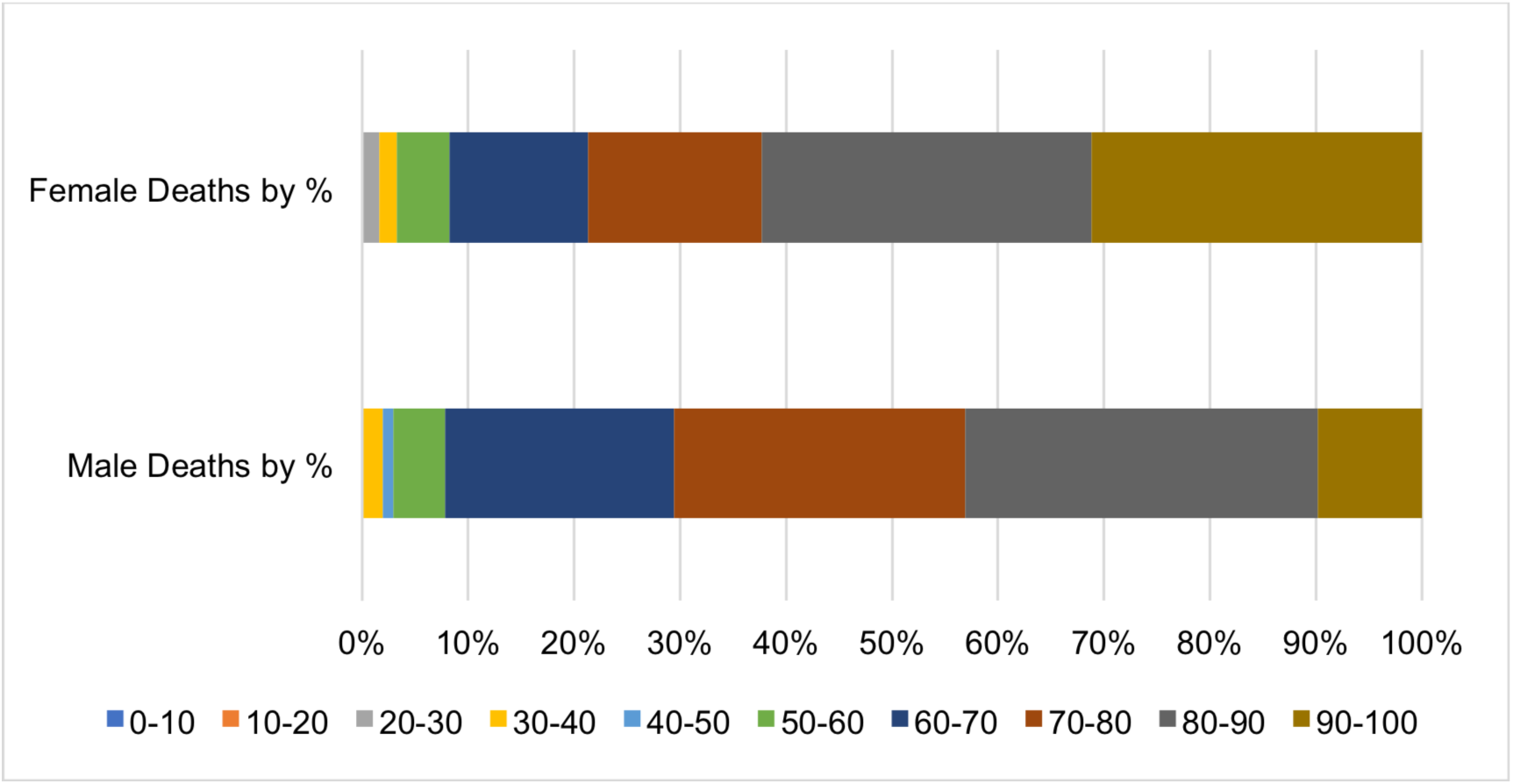

**Figure 4b.**
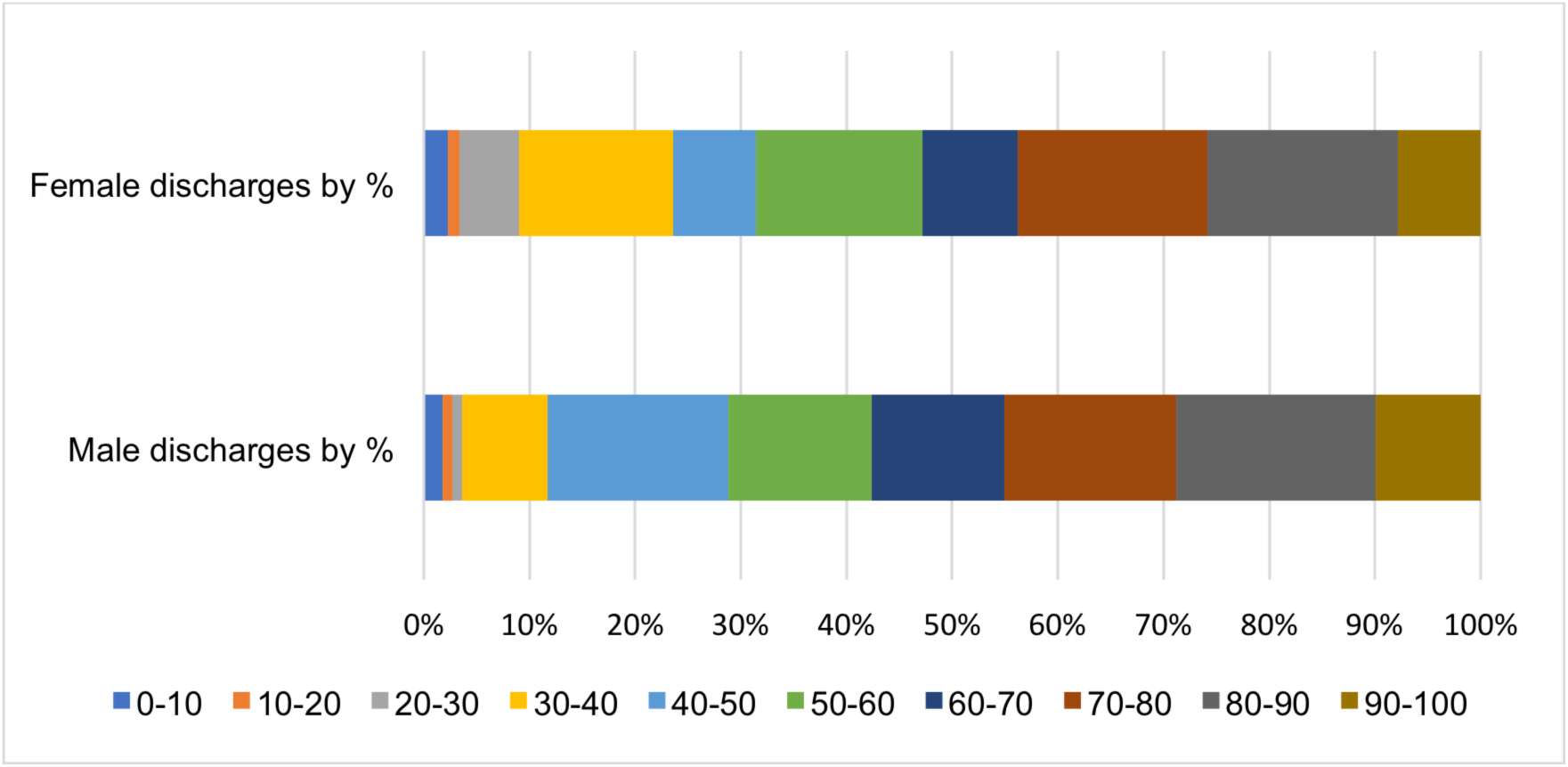

With regard to patients who have been discharged, even though far fewer young patients were hospitalised, when symptoms were severe enough to warrant hospital admission, younger patients were disproportionately more resilient to COVID-19 infection.

When the data is analysed to determine the relative incidence of death against discharge, the plots for male and female patients (Figures 5a&b) suggest that women up to their mid-eigthees are more likely to go home than die. In contrast men admitted to hospital with COVID-19 are more likely to die than survive once they reach their mid-sixtees.

**Figure 5a.**
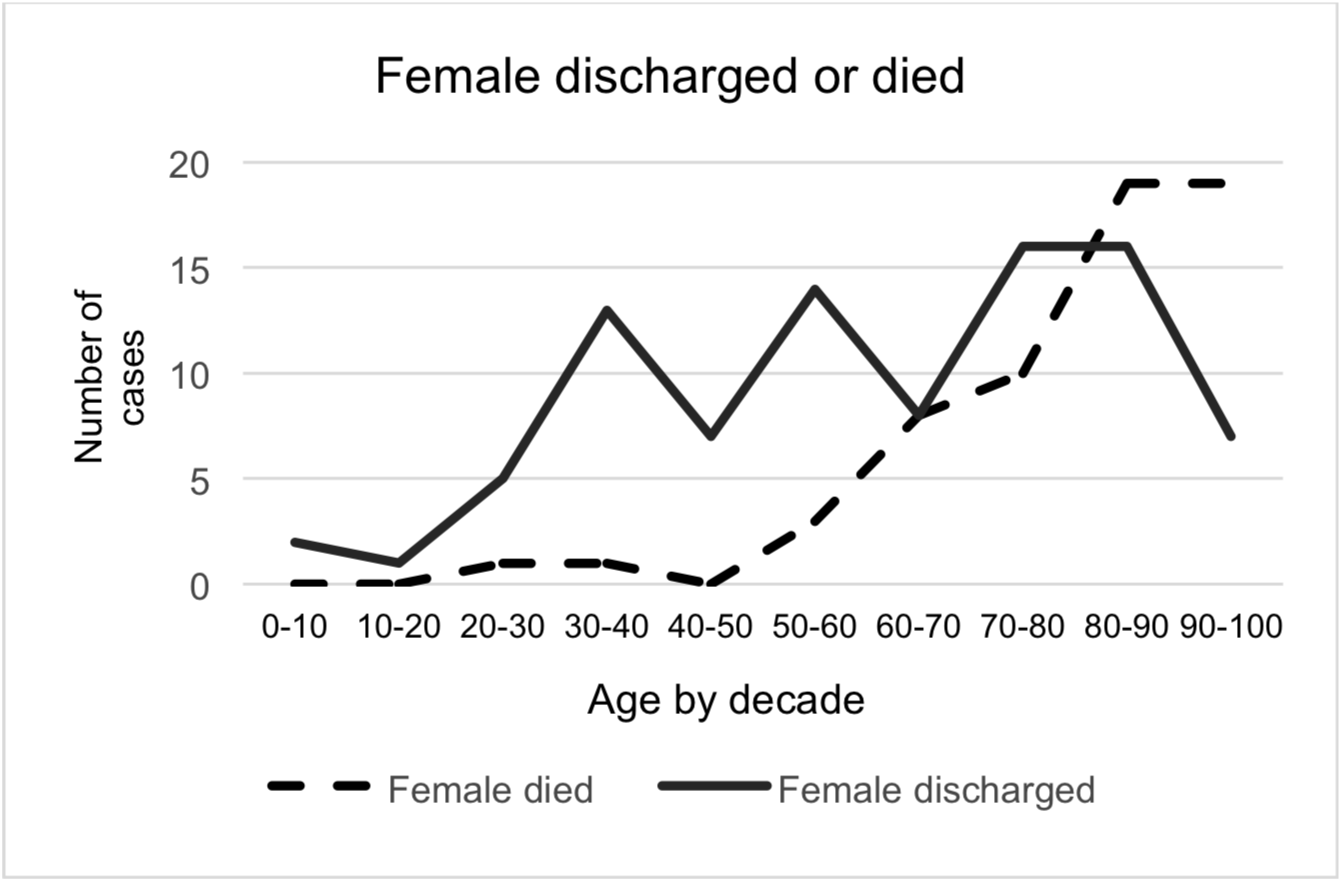

**Figure 5b.**
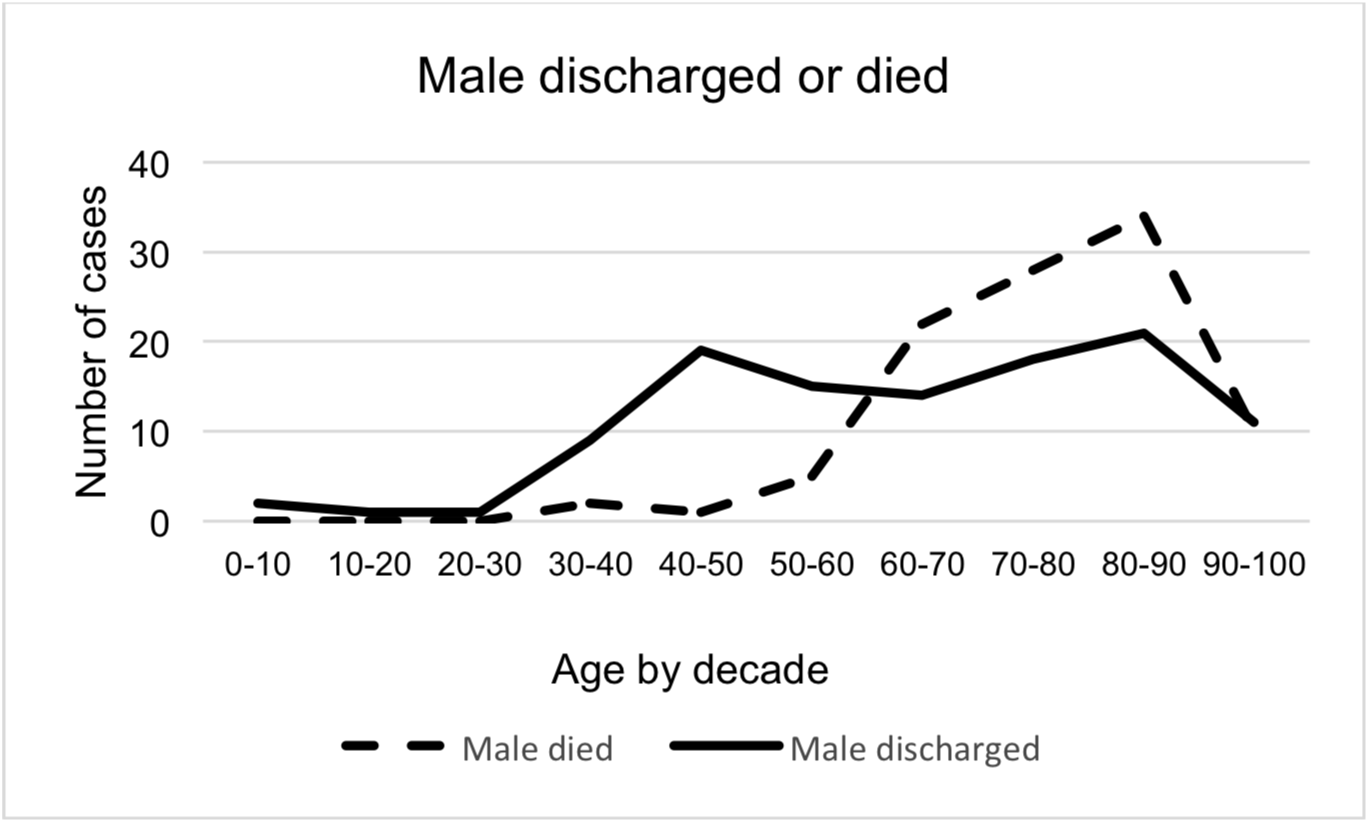

Review of death and discharge rates by Index of Multiple Deprivation Decile was undertaken to see whether the ratio of patients being discharged to those dying differed in the ten deciles. The data did not show any adverse correlation associated with lower Index of Multiple Deprivation Decile groups (Figure 6). Increasing age has already been identified with increasing decile number and other factors may also explain the distribution.

**Figure 6.**
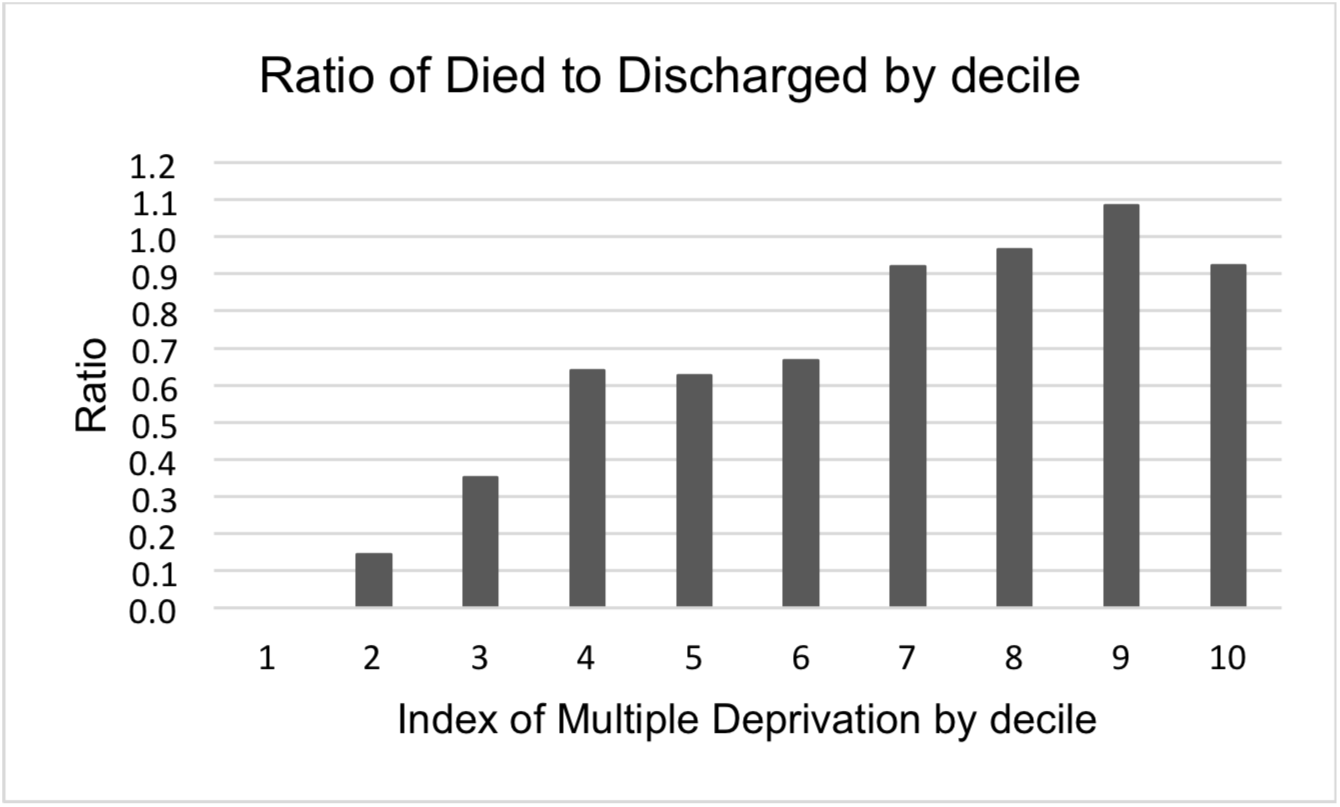

A similar analysis was undertaken with respect to the patients’ ethnicity. The ratio of Died to Discharged was calculated for each group: Unknown Ethnicity 0.41, White 0.93, Mixed race 5.0, Asian 0.59, Black 0.54, Other Ethnicity 0.36. The Died to Discharged ratio of the White group was then compared to each of the other groups to determine whether the differences were statistically significant. The results are shown in Table 4. The two comparisons that showed a statistically significant difference were against the Mixed and Other Ethnic groups.

**Table 4.**

| **Risk ratio**<br>**Died:Discharged** | **Risk Ratio** | **Odds ratio** | **Fisher exact**<br>**P- Value** |
| --- | --- | --- | --- |
| **White:Mixed** | 0.97 | 0.19 | 0.00001 |
| **White:Asian** | 1.05 | 1.57 | 0.1837 |
| **White:Black** | 1.04 | 1.72 | 0.0999 |
| **White: Other ethnic** | 1.05 | 2.55 | 0.0089 |

The surge in admissions to ESTH that started around 15 March 2020, peaked at the beginning of April. The dataset was analysed to ascertain how long the patients remained in hospital after being admitted and tested for COVID-19. Also, whether the length of their hospital admission was affected by the course of their illness. The seven, successive, five-day, cohorts of patients who were tested and found to be positive for COVID-19, between the 6 March 2020 and the 9 April 2020, were reviewed. For each cohort, the number of patients who either died or were discharged (outcome known) was compared against those who remain in hospital (outcome unknown). For this analysis, the date of testing for COVID-19 was used as a surrogate for the time at which clinical suspicion of COVID-19 infection could be inferred. Figure 7a shows, for each time period, the number of patients for whom the outcome was known and the number of patients who remained in hospital on the morning of the 14^th^ of April 2020. Figure 7b shows the percentage of cases in each time-period.

**Figure 7a.**
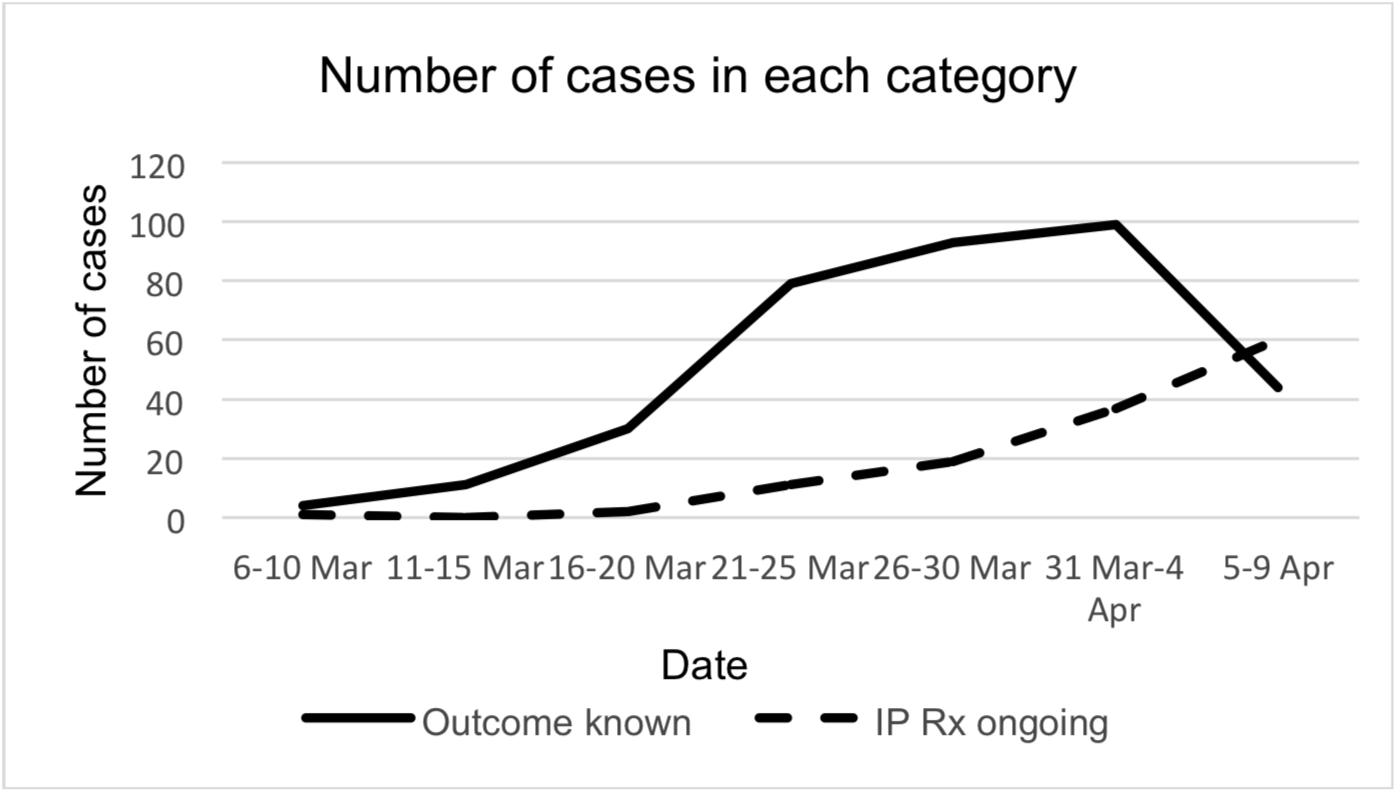

**Figure 7b.**
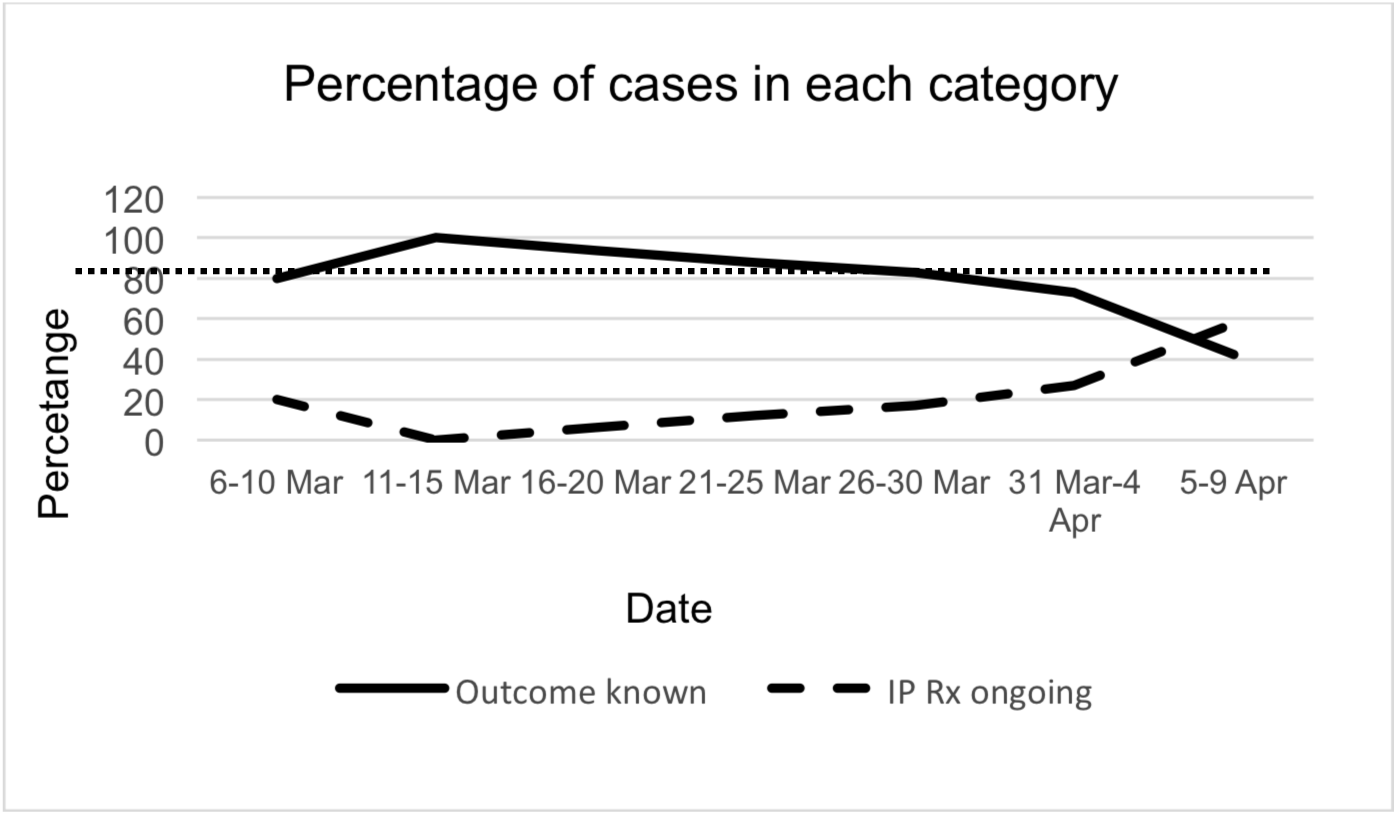

Using the data from the five time-periods with 80% or greater certainty of outcome, (6-10 March, 11-15 March, 16-20 March, 21-25 March and 26-30 March) the interval to death or discharge (LOS) were as detailed in Table 5.

**Table 5.**

|  |  | **From admission** | **From sample** |
| --- | --- | --- | --- |
|  | Average | 11.72 | 6.81 |
| **Intervals to death (days)** | SD | 11.50 | 4.60 |
|  | Min | 0.99 | -0.06 |
|  | Max | 77.13 | 22.00 |
|  | Average | 11.29 | 8.08 |
| **Interval to discharge (days)** | SD | 11.08 | 5.04 |
|  | Min | 0.35 | 0.35 |
|  | Max | 62.78 | 23.48 |

To obtain an indication on how the COVID-19 patients compared with the patients who would have been anticipated if the pandemic had not occurred, we reviewed the emergency admissions to ESTH with a primary diagnosis of respiratory disease, pneumonia or seasonal influenza in the period of 14 March to 14 April 2019.

The demographics and outcome of this cohort are shown in Tables 6a & 6b.

**Table 6a.**

| **Admissions** | **Number** | **Percentage (%)** | **Age** | **SD** | **LOS (Average)** | **LOS (SD)** |
| --- | --- | --- | --- | --- | --- | --- |
| Female | 150 | 51.55 | 61.67 | 30.27 | 8.06 | 12.61 |
| Male | 141 | 48.45 | 58.55 | 32.07 | 8.2 | 13.38 |
| Total | 291 | 100 | 59.9 | 31.13 | 8.13 | 12.96 |

**Table 6b.**
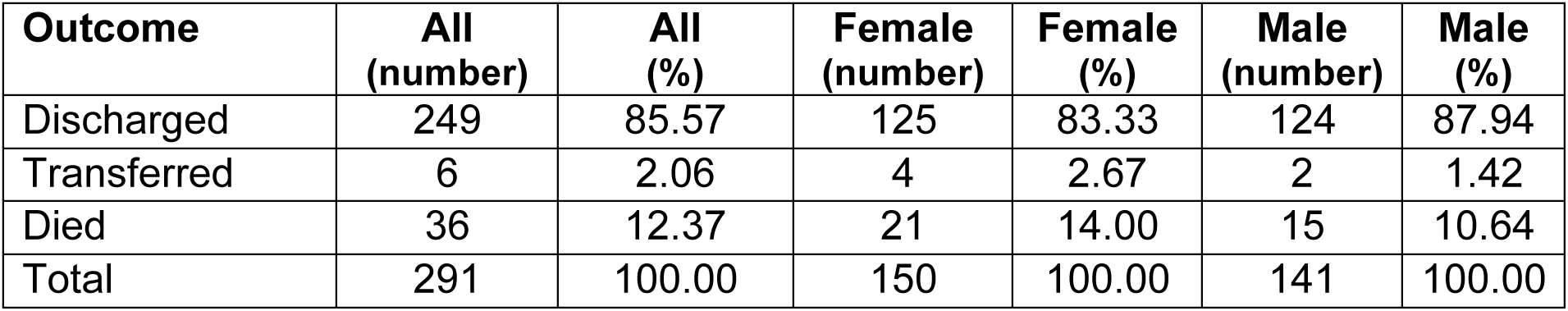

62 of the 500 admissions required ventilator support during their admission. A further six cases were transferred to St Georges and St Thomas’ hospitals. 44 (71%) of the patients ventilated at ESTH were male and 18 (29%) female. The status of the 62 patients by the morning of 14 April is shown in Table 7.

**Table 7.**

| **Outcome** | **Number** | **Percentage of ITU patients (62)** | **Percentage of all patients (500)** |
| --- | --- | --- | --- |
| **Died** | 31 | 50.00 | 6.2 |
| **Still on the ventilator** | 25 | 40.32 | 5 |
| **Stepped down** | 3 | 4.84 | 0.6 |
| **Discharged home** | 3 | 4.84 | 0.6 |
| **Total** | **62** | 100.00 | 12.4 |

When the outcome of the ventilated patients is viewed by age and gender Table 8a & b, no female over the age of 60 has yet left the intensive care unit alive and no male over the age of 50 has left the intensive care unit alive.

**Table 8a.**

| **Females**<br>**Age**<br>**range** | **Died** | **Still**<br>**ventilated** | **Stepped**<br>**down** | **Home** |
| --- | --- | --- | --- | --- |
| **0-10** | 0 | 0 | 0 | 0 |
| **10-20** | 0 | 0 | 0 | 0 |
| **20-30** | 0 | 0 | 0 | 0 |
| **30-40** | 0 | 1 | 1 | 0 |
| **40-50** | 0 | 2 | 0 | 1 |
| **50-60** | 1 | 2 | 2 | 0 |
| **60-70** | 4 | 2 | 0 | 0 |
| **70-80** | 1 | 1 | 0 | 0 |
| **80-90** | 0 | 0 | 0 | 0 |
| **90-100** | 0 | 0 | 0 | 0 |
| **Total** | **6** | **8** | **3** | **1** |

**Table 8b.**

| **Males**<br>**Age**<br>**range** | **Died** | **Still on the**<br>**ventilator** | **Stepped**<br>**down** | **Home** |
| --- | --- | --- | --- | --- |
| **0-10** | 0 | 0 | 0 | 0 |
| **10-20** | 0 | 0 | 0 | 0 |
| **20-30** | 0 | 0 | 0 | 0 |
| **30-40** | 1 | 1 | 0 | 1 |
| **40-50** | 1 | 1 | 0 | 1 |
| **50-60** | 3 | 7 | 0 | 0 |
| **60-70** | 12 | 7 | 0 | 0 |
| **70-80** | 7 | 1 | 0 | 0 |
| **80-90** | 1 | 0 | 0 | 0 |
| **90-100** | 0 | 0 | 0 | 0 |
| **Total** | **25** | **17** | **0** | **2** |

Analysis of duration of ventilation by outcome is shown in Table 9.

**Table 9.**

| **Outcome** | **Mean days on ventilator** | **SD** | **Minimum days on ventilator** | **Maximum days on ventilator** | **Range (days)** |
| --- | --- | --- | --- | --- | --- |
| **Still ventilated** | 11.85 | 8.11 | 0 | 36 | 0 - 36 |
| **Improved & off ventilator** | 7.75 | 3.69 | 3 | 12 | 3 - 12 |
| **Died** | 6.74 | 4.34 | 0 | 18 | 0 - 18 |

## Discussion

Identification of the first 500 admissions was complicated by several factors. Some of the patients had come into hospital for unrelated problems and developed COVID-19 symptoms during their time as an in-patient. This is reflected in the longer mean interval from admission to testing (26.37 days) for the first 51 admissions against the mean interval of 1.37 days for the subsequent 449 patients. Also, 20 of the patients had repeat admissions recorded over the three-month period. In these cases, we used the date of the most recent admission for our calculations. However, this approach is also imperfect as some of the cases were re-admitted for problems unrelated to COVID-19 infection and others were readmitted after the sample that proved positive had been taken during a first admission and the patient had been discharged prior to the result becoming available.

This service evaluation provides a preliminary analysis based on a very small subset of the information that is being collected locally and nationally on COVID-19 patients. The conclusions that can be drawn from such a simplistic snapshot are very limited and the picture that our data provides is inevitably distorted by the many confounding factors that we have not assessed. The data used for this service evaluation was obtained from the Trust’s Information Management team as a subset of the information available from the ESTH Clinical Manager and Ward Watcher systems and a subset of the data recorded by the intensive care team. We did not review any in-patient notes or extract any in-patient vital-sign recordings, laboratory results or medications prescribed during the admissions from the Trust’s electronic data repositories.

The dataset lacks any clinical information such as Body Mass Index (BMI), ongoing and previous medical conditions, recent and repeat medications history, immunisation history, allergies, or other pre-existing vulnerabilities. The latter information is recorded on the General Practitioners’ ‘Summary Care Record’ and all UK, NHS, General Practitioners. This information is now recorded electronically on one of four systems (EMIS Web, SystmOne, Vision Care and Microtest Evolution). To provide a thorough analysis of the patients and their treatment journey would necessitate inclusion of information from all primary and secondary care data repositories. However, in the ESTH Trust, other information such as the A&E admission data, vital signs recorded during intensive care treatment, drugs administered during intensive care treatment and ventilation data is all recorded on paper and not available from any electronic data repository. A hybrid data capture system that combines primary and secondary care electronic data repositories with clinical data recorded on paper would be required to compile a complete composite of the patients and their treatment.

It is impossible to specify that the surge in admissions started on a particular day. Nevertheless, this was within a day or two of the 15 March 2020. The UK wide lockdown started 8 days later and the peak of admissions to ESTH occurred after a further 7-9 days. This would be consistent with a interval between being infected and requiring hospital admission of 6-8 days.

56.8% of the admissions were male. This is lower than the 62.6% reported by Petrilli et al [7] in their analysis of 1999 patients admitted for in-patient, COVID-19 treatment at a New York hospital between 1 March 2020 and 2 April 2020 and the 68% reported by Chen et al in their study of 99 patients admitted to Wuhan Jinyintan Hospital from 1 January to 20 January 2020. The average age of ESTH patients was higher 69.32 years (SD19.23 years) than either the US report mean 62 years or the Chinese study mean 55.5 years. The increased incidence with increasing age is consistent with previous reports. However, previous reports have not reviewed the effect of age by decade or considered social deprivation.

A novel aspect of this study is the analysis of likelihood of survival for the different age groups. This has revealed that males and females under the age of 50 can expect to recover. Females remain more likely to leave hospital alive until they reach their late 70’s while males are more likely to die than leave hospital alive after their mid 50’s.

Our review of a patient’s Index of Multiple Deprivation did not reveal any adverse finding associated with the greater deprivation of the lower deciles but was limited in that the population of patients admitted were mostly from more affluent sections of the community.

Our analysis of Ethnicity suggests that the only subgroup more likely to die than the White subgroups are the ‘Mixed Races’. However, the number of individuals in this subgroup is very small (5 cases) and no confounding factors, such as age, BMI or comorbidities have been considered. We suspect that this result is erroneous. Nevertheless, the lack of evidence of poorer outcome for the ethnic minorities and statistically significant finding of better outcomes for the ‘Other Ethnic’ group is at variance with a widely publicised recent US media report [8]. This may reflect a difference in social and healthcare provision between the UK and US.

Our review of the outcome of in-patient treatment indicates that 80% of patients will have died or been discharged within three weeks. Furthermore, the mean and standard deviation figures for the two outcomes; mean 11.72 days to death (SD 6.81) and mean 11.29 days to discharge (SD 8.08) are remarkably similar. This translates to less than 16% of outcomes being unknown by three weeks. If the south London surge did peak at the beginning of April, the need for hospital and Intensive care facilities will lessen from mid-April.

The COVID-19 patients differed in several ways from the cohort who were admitted during the same period in 2019. The gender and mortality ratios were reversed and the mean age of the COVID-19 patients was a decade higher (69.32 against 59.9 years). The mortality rate of the COVID-19 patients was approximately 2.5 times higher (32.6% against 12.27%) and the mean duration of their admissions was three days longer than the 2019 cohort. In these respects, the COVID-19 infection is proving to be a more challenging medical problem, particularly for elderly men.

With regard to patients who have required ventilator support, the data obtained at ESTH is trivial in comparison to the national experience that is being reported by the ‘*ICNARC*’ study group [9]. The ESTH experience represents approximately 1% of the total *ICNARC* experience and should be interpreted with caution. Nevertheless, the ESTH data is consistent with national outcomes and if the outcome for patients requiring prolonged ventilation does not prove any more successful than seen in the first 11 days, staff morale may be compromised and public support for the nation’s investment in critical care for COVID-19 patients may be challenged.

While the duration of the COVID-19 pandemic remains unknown, detailed data collection and an infrastructure to provide this service remains a priority that will help care providers make the best decisions for the unfortunate individuals who develop more severe symptoms. It will also help the Government to allocate resources to fight the pandemic most effectively and with least long-term damage to society. When the restrictions of the current lockdown are lifted, it will be the younger members of society who will be at least risk of developing life-threatening symptoms, should they become infected.

## Data Availability

The data that support the findings of this study are available from the corresponding author, REF upon reasonable request. The data is not publicly available.

## Acknowledgments

The authors of this publication would like to thank all the members of the Outcomes team at the South West London Elective Orthopaedic Centre, Mr Andrew Dodds and the Information Management team at Epsom and St Helier University NHS Trust for all their hard work in supporting this work. With special thanks to Professor D Kader, Professor D Sochart, Mr V Asopa, Mr P Mitchell and Mr R Twyman for their support and guidance throughout the collection of data and preparation of this work.

